# Shannon Entropy of Gray Matter Eigenmodes: A Novel Biomarker for Alzheimer’s Disease and Heterogeneous MCI Trajectories

**DOI:** 10.1101/2025.06.12.25329476

**Authors:** Yumeng Li, Gaoping Long, Xinyue Zhang, Xin Li, Zhanjun Zhang

## Abstract

**Background:** Current Alzheimer’s disease (AD) diagnostics rely on late-stage cognitive assessments or invasive biomarkers. Neuroimaging offers non-invasive alternatives, but single-modality approaches (structural atrophy or functional connectivity) face limitations in sensitivity and specificity for early detection. We introduce entropy and temperature, novel structure-function coupling (SFC) biomarkers based on gray matter eigenmodes, to quantify cortical disorganization in early AD.

**Methods:** We analyzed multimodal MRI (structural + resting-state fMRI) and amyloid-PET data from two independent cohorts (BABRI: N=135; ADNI: N=275), including cognitively normal (CN), mild cognitive impairment (MCI), and AD participants. We computed Shannon entropy by projecting fMRI time series onto individual structural eigenmodes, and temperature based on eigenmode-based reconstruction of functional connectivity. These indexes were evaluated for diagnostic classification (SVM), prediction of amyloid- β (A β) burden (regression), and stratification of MCI subtypes (reversed/stable/progressed).

**Results:** Entropy was significantly elevated in AD compared to CN and MCI participants across both cohorts (Δ=8-21%, p<0.001), with left-hemisphere entropy showing superior diagnostic accuracy (AUC=0.901 for CN vs. MCI; AUC=0.873 for CN vs. AD). Right-hemisphere and global entropy robustly predicted Aβ deposition (error reduction: 38.7–42.1% vs. baseline, p<0.01). Entropy stratified MCI into distinct subtypes: progressors (MCI→AD) showed higher entropy and Aβ than stable/reverted MCI (p<0.001) and exhibited a biphasic entropy trajectory during conversion. Temperature indices did not show significant differences across diagnostic groups.

**Discussion:** Entropy derived from gray matter eigenmodes emerges as a sensitive and dual-purpose biomarker for AD diagnosis and pathological prediction. Its hemispheric asymmetry (left: optimal classification; right: Aβ prediction) and ability to detect nonlinear MCI progression offer mechanistic insights for early intervention.

## 1 Introduction

Alzheimer’s disease (AD), a highly prevalent neurodegenerative disorder, is known to begin its pathological progression 10 to 20 years before the onset of clinical symptoms[1]. However, current diagnostic approaches primarily rely on late-stage cognitive assessments and invasive cerebrospinal fluid (CSF) tests, which are limited by their invasiveness, high cost, and limited accessibility[2]. Therefore, the development of non-invasive, highly sensitive early screening techniques is crucial for identifying a precise intervention window for effective treatment of the disease[3].

Neuroimaging has emerged as a promising non-invasive biomarker for the early detection of Alzheimer’s disease (AD) and cognitive impairment[4]. Structural features such as hippocampal gray matter volume and medial temporal lobe atrophy have shown diagnostic value in imaging studies[5, 6]. However, their practical application still faces several limitations. First, changes in individual gray matter volume and cortical thickness exhibit considerable inter-individual variability[7, 8]. These features are influenced by a range of genetic, environmental, and lifestyle factors. Although recent research advocates for multidimensional models that integrate genetic and environmental variables to assess early disease susceptibility[9], such comprehensive monitoring remains impractical for widespread use in the short term. Due to this variability and overlap, relying on a single gray matter feature may result in high false-positive rates (misclassifying healthy individuals with naturally lower gray matter or faster aging as high risk) or false negatives (missing high-risk individuals with early pathology but preserved gray matter) [10]. Differentiating between “normal aging” and “pathological” atrophy is particularly challenging during the early disease stages[11, 12]. Second, in the earliest detectable phases of AD—such as preclinical AD or early MCI—structural changes in gray matter may be subtle and focal[13, 14]. These early alterations are often too minor to be reliably captured by structural MRI alone. As a result, using gray matter changes as early indicators may lead to missed intervention windows. Moreover, gray matter atrophy progresses relatively slowly, and a single time-point measurement cannot capture the dynamic speed of disease progression[15, 16]. Resting-state functional MRI (rs-fMRI), another key neuroimaging modality, can detect brain network dysfunction, which may precede structural atrophy. It has shown great potential for early screening of individuals at risk for AD and MCI[17–19]. However, like structural markers, functional connectivity also suffers from high variability and temporal lag when used as a single diagnostic feature.[20] Additionally, brain network data are extremely high-dimensional, involving thousands of connections[21]. Identifying disease-relevant features requires a large number of statistical comparisons, increasing the risk of false positives. Furthermore, many functional connectivity alterations lack spatial specificity for AD and are difficult to replicate across studies due to methodological inconsistencies and limited standardization[22].

While multi-modal neuroimaging methods have been proposed to overcome the limitations of the single-modal for early Alzheimer’s disease (AD) detection[23–25]. More specifically, these shortcomings can be explained in two perspectives. First, most studies rely on a simplistic "feature stacking" strategy, where structural features and functional connectivity are independently extracted and subsequently fused. Crucially, this post-hoc combination fails to capture the inherent, dynamic interdependencies between brain structure and function that are likely central to pathological processes. Second, the dominant paradigm concentrates on modeling associations between white matter connectivity and resting-state functional connectivity[26, 27]. In fact, this white-matter-centric SFC framework faces some key challenges for early AD screening, which can be listed as follows.

a. White matter connectomes often show low accuracy in predicting functional connectivity patterns[28]. Thus, this white-matter-centric SFC cannot provide a mechanism to explain the abnormal SFC in early AD. Essentially, there is considerable functional redundancy not constrained by the specific white matter pathways being measured.
b. White matter connectivity primarily describes long-range, inter-regional communication pathways. Consequently, it overlooks the critical physical constraints imposed by gray matter microstructure and geometry at the local microcircuit level.[29, 30] This omission is particularly detrimental, since accumulating evidence suggests that early AD pathology (e.g., molecular disruptions, astrocytic dysfunction) begins in the local gray matter microenvironment[31, 32]. These changes occur well before widespread white matter degeneration or large-scale network failure becomes apparent[33].

The shortcomings of previous researches on structure–function coupling (SFC) suggest us to focus on the dynamic interactions between gray matter structure and brain function. A widely adopted approach in this direction is the eigenmode method, by which the eigenmodes determined by the structural information is used to analyze the brain function[34–36]. Recent work published in Nature has demonstrated that eigenmodes derived from cortical geometry—not white matter connectivity—can effectively explain and predict large-scale functional brain activity[30]. This challenges the long-standing paradigm that “white matter connectome determines function,” suggesting instead that the geometry of gray matter imposes a more fundamental constraint on functional organization. Moreover, according to current models of early Alzheimer’s disease (AD) pathogenesis, molecular abnormalities initially disrupt the local gray matter microenvironment, such as through dysfunction of astrocytic water channels and ionic imbalance[37–40]. Therefore, we propose that gray matter eigenmodes, which serve as constraints on brain function, may capture subtle alterations associated with the progression of AD better.

Building upon this paradigm shift, we introduce two novel Structural-Functional Coupling (SFC) indexes based on gray matter eigenmodes in this article. More explicitly, the eigenmodes derived from individual high-resolution structural MRI (sMRI) are treated as an intrinsic spatial basis for functional activation. To capture the characteristics of the brain functions supported by the structural eigenmodes, we propose two neurophysiologically meaningful indexes. The first index is the entropy of the distribution of the brain function over the eigenmodes. Specifically, by projecting resting-state fMRI time series onto the gray matter eigenmode basis to compute activation coefficient for each mode, the Shannon entropy of these coefficients quantifies the dispersion of functional activity across eigenmodes. In other words, lower entropy values indicate that activity is concentrated in fewer modes, which reflects a more ordered state; Higher entropy values indicate functional dispersion across a broader range on eigenmodes, which reflects a more disordered and distributed activation profile. The second index is the temperature of the reconstructing whole-brain functional connectivity matrix based on the gray matter eigenmodes. Specifically, we assess the predictive accuracy of this reconstruction across eigenmodes ranked by frequency, and fit it to a cumulative distribution function of exponential decay distribution. Then, the temperature is given by the exponential decay coefficient. A steeper decay (lower temperature) suggests that low-frequency modes dominate the functional architecture, while a flatter decay (higher temperature) indicates greater contributions from high-frequency modes. Together, these indexes offer a principled and interpretable framework for quantifying the structure-function relationship grounded in cortical geometry, thereby advancing a geometry-informed model of brain dynamics.

In this study, we provide preliminary validation of entropy- and temperature-based indexes for distinguishing between normal and pathological aging. We hypothesize that individuals with pathological aging will exhibit significantly greater disorder in brain system dynamics, and that these indexes offer enhanced sensitivity for detecting preclinical Alzheimer’s disease (AD) along with more specific mechanistic insight. In particular, we aim to establish Shannon entropy as a novel, non-invasive biomarker for tracking AD progression, given its responsiveness to cortical network disorganization beyond conventional structural or functional measures. To further address the heterogeneity of mild cognitive impairment (MCI), we stratify clinically defined MCI cases into reversed, stable, and progressed subtypes based on longitudinal entropy trajectories. This framework is applied across two multimodal cohorts (BABRI, N=135; ADNI, N=275), using entropy and temperature indexes to evaluate diagnostic specificity, characterize disease progression, and predict amyloid-β (Aβ) burden, with machine learning and regression models optimized for classification and biomarker associations.

## 2 Methods

### 2.1 Participants

The participants were from the Beijing Aging Brain Rejuvenation Initiative (BABRI) [41] and the Alzheimer’s Disease Neuroimaging Initiative (ADNI) [42]. All participants were classified as NC or MCI or AD at baseline. All the participants completed florbetapir PET, structural MRI and resting functional MRI. Briefly, the diagnostic criteria for MCI included subjective memory complaints, impairment in at least one cognitive domain (1.5 standard deviations or more), relatively preserved general cognitive function, and intact ability to perform activities of daily living [43]. The criteria for normal cognition were no cognitive complaints, a Mini-Mental State Examination (MMSE) score of no less than 24, and being able to perform the normal activities of daily life. AD was diagnosed according to the criteria of the National Institute of Neurological and Communicative Disorders and Stroke and the Alzheimer’s Disease and Related Disorders Association Dementia [44, 45] and further evaluated by brain CT or MRI. A total of 135 eligible participants from the BABRI and a total of 360 eligible participants from the ADNI were included in this study.

In the data derived from ADNI, we also incorporated the longitudinal follow-up information of the MCI group. All participants in the MCI follow-up cohort underwent structural MRI and resting-state functional MRI examinations. A total of 49 eligible participants were ultimately enrolled. Among them, 10 subjects were classified as reversed MCI (reverting from MCI to cognitively normal (CN) during follow-up), 26 maintained stable MCI diagnoses, and 13 progressed to Alzheimer’s disease (AD) (transitioning from MCI to AD).

The study was conducted in accordance with the institutional review board (IRB) at the Imaging Center for Brain Research at Beijing Normal University (protocol code ICBIR_A_0041_002_02 and date of approval 03.2015). All participants provided written informed consent for our protocol, which was approved by the ethics committee of the State Key Laboratory of Cognitive Neuroscience and Learning, Beijing Normal University.

### 2.2 MRI data acquisition and processing

High-resolution T1-weighted MRI data were collected from each participant from the BABRI and ADNI using either 1.5-T scanners (participants from ADNI-1) or 3-T scanners (participants from BABRI and ADNI-GO&2); the acquisition parameters for each study have been published previously [46, 47].

### 2.3 MRI image acquisition and data processing

For all subjects, cortical reconstruction of T1-weighted images was performed using FreeSurfer version 5.3 (http://surf er.nmr.mgh.harvard.edu)[48]. This process involved registration to a template, intensity normalization, gray/white matter segmentation, and tessellation of gray/CSF and white/gray boundaries. Cortical surfaces were inflated and normalized via spherical registration, with cortical thickness defined as the shortest distance between the pial and white matter surfaces. The average regional cortical thickness was calculated without manual correction. The procedure included bias field correction, intensity normalization, and skull stripping using a watershed algorithm, followed by white matter segmentation, surface definition, and topology correction of the reconstructed surfaces. We used a triangular surface mesh representation of the midthickness human cortical surface. This representation, comprising 32,492 vertices in each hemisphere, was obtained from a down sampled, left–right symmetric version of the FreeSurfer’s fsaverage population-averaged template

(https://github.com/ThomasYeoLab/CBIG/tree/master/data/templates/surface/fs_LR_164k). Note that the template is independent of the data sample used in our analyses, thus avoiding concerns about circularity.

### 2.4 The preprocessing pipeline for functional neuroimaging data

Resting-state fMRI data were preprocessed using fMRIPrep (https://fmriprep.org/, V20.1.3), a Python-based automated tool integrating advanced neuroimaging packages like FSL, ANTs, FreeSurfer, and AFNI, designed for optimized fMRI preprocessing [49].

The procedure included head motion estimation, slice - timing correction, fMRI-to-T1w registration, fMRI-to-MNI152 standard - space normalization, confound estimation, and regression. Specifically, head motion parameters were estimated and saved for subsequent regression. Slice-timing correction adjusted for differences in slice acquisition times. The fMRI data were coregistered to the T1 - weighted structural image for precise alignment and then normalized to the MNI152 space for group - level analysis. Confounds such as head motion parameters, white matter, and CSF signals were estimated for regression. Post - preprocessing steps comprised linear drift removal, Gaussian smoothing (6 mm FWHM), and nonlinear band - pass filtering (0.01 - 0.1 Hz). Quality control involved visually checking T1w images and excluding subjects with excessive head motion (maximum head motion > 3 mm or 3°, or mean FD > 0.5 mm). All analyses were conducted at the voxel level to ensure accuracy and consistency.

### 2.5 Spatiotemporal Mode Decomposition of Neural Dynamics Through Structural Eigenmodes

The computational pipeline for investigating the coupling between cortical geometry and functional activity was implemented through four sequential modules (Fig. X), with all analyses conducted in individual cortical space to preserve neuroanatomical specificity.

#### Structural Eigenmode Computation

Cortical surface reconstruction was performed using FreeSurfer v7.2 (recon-all pipeline). Laplace-Beltrami Operator (LBO) was defined based on the metric describing the midthickness surfaces geometry. Specifically, Mid-thickness surfaces were converted to VTK format, and the Laplace-Beltrami Operator (LBO) was given through custom Python scripts [30]. Eigenmodes were computed using the eigenvalue equation:

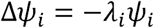

where Δ denotes the LBO, *ψ*_*i*_ represents the *i*-th eigenmode, and *λ_i_* is the corresponding eigenvalue. This computation was generating frequency-ranked cortical vibration modes.

### Functional Data Processing

Firstly, FreeSurfer’s mri_vol2surf (v7.2) implemented grayordinate-wise projection through ray-casting sampling (256 iterations, 0.1 mm step size) onto the native space, preserving cortical columnar organization. Secondly, vertex-level temporal dynamics were encoded in sparse matrices (N = vertices × M TRs), which maintained native temporal resolution. This processing chain ensured topological correspondence between structural eigenmodes and functional time series across spatial scales.

### Cortical Mask Generation

Individual-specific cortical masks were generated through a three-stage surface-constrained protocol. First, probabilistic tissue segmentation from FreeSurfer’s aparc.aseg.mgz output was thresholded at 50% confidence level to minimize partial volume effects. Subsequently, a binarization procedure assigned value 1 to vertices within anatomically verified cortical regions (Desikan-Killiany atlas) and 0 to non-cortical structures (subcortical nuclei, white matter, ventricles). Finally, the volumetric mask was projected onto native cortical surfaces, generating exclusion masks that systematically removed subcortical and ventricular components while preserving cortical geometry in individual surface space. This process ensured topological consistency between structural and functional data representations.

### Computational Implementation

All pipelines were executed on HCP-style processing clusters using Python 3.8 and Bash scripting. Surface operations employed FreeSurfer v 5.3 and Connectome Workbench v1.5 commands, with quality assurance via visual inspection of surface-normal vectors and Euler characteristics.

2.6 Calculation of temperature and Shannon’s entropy Cortical eigenmodes (200 modes) and resting-state fMRI time series (N vertices × M TRs) were first aligned in individual surface space using FreeSurfer-processed mid-thickness meshes. Shannon’s entropy H was computed via:

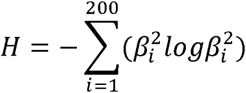

where *β_i_* were obtained by projecting the time-averaged fMRI data onto each eigenmode basis functions. Temperature estimation is given by a fitting of functional connectivity (FC) reconstruction accuracy:

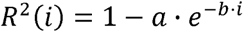

where *R*^2^(*i*) o represents the cumulative reconstruction accuracy of functional connectivity (FC) using the first i structural eigenmodes and *R*^2^(*i*) was fitted to a cumulative distribution function of exponential distribution. The characteristic temperature parameter *T* was defined as *T= 1/b* through nonlinear least-squares optimization (MATLAB fminsearch, initial guess [a=1, b=0], convergence tolerance 1^e-4^). FC matrices were parcellated using Schaefer-400 atlas[50], with upper-triangular elements correlated against empirical FC to compute reconstruction accuracy. All computations were constrained to cortical vertices using individually masked surfaces, ensuring exclusion of subcortical artifacts.

### 2.7 Statistical Analysis

To compare demographic characteristics (analyzed via chi-square test for sex distribution), temperature, and entropy indexes across groups, an analysis of covariance (ANCOVA) was performed with age, education, and sex as covariates. Partial correlation analyses were conducted to assess the relationship between Aβ accumulation and entropy indexes across all participants, adjusting for the aforementioned covariates. Spearman correlation was employed to evaluate associations between disease progression stages and both entropy and temperature indexes.

#### Longitudinal Rate of Change Analysis

For each participant, the rate of entropy change between consecutive follow-ups was calculated as (Entropy_t+1_ – Entropy_t_)/Time. Paired t-tests were conducted to compare these rates across the two intervals within each subgroup (reversed MCI, stable MCI, and progressed MCI), aiming to identify non-linear trends in entropy and temperature dynamics during disease progression.

#### Multi-Class SVM Classification

A multi-class support vector machine (SVM) model with a "one-vs-all" strategy was implemented to evaluate the classification efficacy of neuroimaging indexes (left-hemisphere entropy, right-hemisphere entropy, global entropy, and Aβ deposition) across cognitive states (CN, MCI, AD). Leave-one-out cross-validation (LOOCV) was employed: for each subgroup comparison (CN vs. MCI, CN vs. AD, MCI vs. AD), one sample was iteratively excluded as the test set, while the remaining samples were used to train the model. Model performance was assessed via sensitivity, specificity, and AUC values, with emphasis on hemispheric entropy asymmetry and Aβ pathology in predicting early-stage transitions.

#### Prediction of Aβ Deposition

Under the LOOCV framework, linear regression models were constructed to predict standardized Aβ deposition values using individual entropy indexes as predictors. For each indexes, a generalized linear regression model was trained on all but one sample, and the root mean square error (RMSE) was computed to evaluate prediction accuracy. A baseline model predicting Aβ using the sample mean was established, and effective predictors were identified as those with RMSE significantly lower than the baseline (ΔRMSE < 0, *P*_permutation_ < 0.05).

## 3 Results

### 3.1 Participant Characteristics

The clinical characteristics of participants from both the BABRI (N = 135) and ADNI (N = 275) cohorts revealed distinct demographic and cognitive profiles across diagnostic groups (See Table 1). In the BABRI cohort, no significant differences were observed in sex distribution (χ² = 3.32, *p* = 0.19), age (*F* = 0.51, *p* = 0.61), or education years (*F* = 1.95, *p* = 0.15) among groups, whereas marked cognitive stratification emerged in Mini-Mental State Examination (*F* = 152.3, *p* < 0.001) and Auditory Verbal Learning Test (*F* = 48.5, *p* < 0.001). Similarly, the ADNI cohort showed nonsignificant sex (χ² = 1.21, *p* = 0.55) and age differences (*F* = 2.14, *p* = 0.12), though education years exhibited marginal variation (*F* = 3.86, *p* = 0.02), with pronounced cognitive decline in MMSE (*F* = 236.2, *p* < 0.001) and AVLT (*F* = 98.8, *p* < 0.001). Both cohorts demonstrating progressive cognitive deterioration from NC to AD (all pairwise *p* < 0.05, Bonferroni-corrected), underscoring diagnostic validity while reflecting cohort-specific demographic influences. Given the significant between-group differences in education levels within the ADNI cohort, all subsequent analyses utilized residualized variables adjusted for demographic confounders (age, sex, and education years) through linear regression modeling to mitigate potential confounding effects.

**Table 1.** Characteristics and neuropsychologic test results

| | Total | NC | MCI | AD | F/$\chi^2$ | P |
| --- | --- | --- | --- | --- | --- | --- |
|  | n = 135 | n = 76 | n = 30 | n = 29 |  |  |
| **BABRI sample (N = 135)** |  |  |  |  |  |  |
| Sex (female, %) | 79 (58.5%) | 43 (56.6%) | 15 (50%) | 21 (72.4%) | 3.32 | 0.19 |
| Age (years) | 68.23 ± 8.09 | 67.08 ± 7.63 | 68.73 ± 8.35 | 70.72 ± 8.65 | 0.51 | 0.61 |
| Education (years) | 12.60 ± 3.80 | 13.19 ± 3.56 | 12.07 ± 3.35 | 11.62 ± 4.62 | 1.95 | 0.15 |
| Aβ accumulation | 0.99±0.12 | 0.95±0.11 | 1.00±0.11 | 1.08±0.13 | 13.14 | < 0.001<sup>1,2,3</sup> |
| MMSE | 24.11 ± 6.96 | 27.58 ± 1.65 | 26.30 ± 1.56 | 12.76 ± 7.11 | 152.3 | < 0.001<sup>1,2,3</sup> |
| AVLT (N1-N5) | 21.63 ± 12.20 | 29.08 ± 8.90 | 16.90 ± 8.15 | 6.46 ± 4.68 | 48.5 | < 0.001<sup>1,2,3</sup> |
| **ADNI sample (N = 275)** |  |  |  |  |  |  |
| Sex (female, %) | 140 (50.9%) | 59 (55.1%) | 60 (48%) | 21 (50%) | 1.21 | 0.55 |
| Age (years) | 72.53±6.99 | 72.27±6.55 | 72.06±7.30 | 74.56±6.95 | 2.14 | 0.12 |
| Education (years) | 16.24±2.66 | 16.82±2.37 | 15.97±2.87 | 15.57±2.47 | 3.86 | **0.02** |
| Aβ accumulation | 27.49±2.83 | 28.97±1.26 | 27.97±1.79 | 22.31±2.37 | 37.112 | < 0.001<sup>1,2,3</sup> |
| MMSE | 27.49±2.83 | 28.97±1.26 | 27.97±1.79 | 22.31±2.37 | 236.2 | < 0.001<sup>1,2,3</sup> |
| AVLT (N1-N5) | 38.04±12.50 | 46.08±9.80 | 36.53±10.16 | 22.02±6.43 | 98.8 | < 0.001<sup>1,2,3</sup> |

Note: The measured data are represented by the mean and standard deviation. The p value for sex was obtained using a Chi-square test. AVLT: Auditory Verbal Learning Test; $p^1$ indicates a significant difference between the NC and MCI groups. $p^2$ indicates a significant difference between the NC and AD groups. $p^3$ indicates a significant difference between the MCI and AD groups, $p < 0.05$ , the same as below.

### 3.2 Entropy and Temperature Profiles Across Diagnostic Groups

#### BABRI Cohort (N = 135)

Significant intergroup differences emerged in entropy measures but not in temperature indexes. For **entropy in left hemisphere**, ANCOVA revealed moderate group differences (F=3.07, *p*<0.05), with post hoc comparisons indicating AD exhibited higher entropy than both NC (*p*<0.05) and MCI (p<0.01). **Entropy in right hemisphere** demonstrated stronger discrimination (F=7.64, *p*<0.001), where AD > NC (*p*<0.001) and AD > MCI (*p*<0.01). Global entropy (**entropy**) showed intermediate effects (F=4.1, *p*<0.01), with AD exceeding NC (*p*<0.05) and MCI (*p*<0.01). In contrast, temperature parameters showed no significant group differences (all F<13.14, *p*>0.05), suggesting temperature indexes lack diagnostic specificity in this cohort.

#### ADNI Cohort (N=275)

Entropy measures showed robust diagnostic stratification. **Entropy in left hemisphere** (F=10.77, *p*<0.001) differentiated AD from NC (*p*<0.001) and MCI (p < 0.01). Similarly, **entropy in right hemisphere** (F = 12.08, *p*<0.001) and global **entropy** (F = 12.21, *p*<0.001) exhibited AD > NC (*p* < 0.001) and AD > MCI (*p* < 0.001). Temperature indexes again showed no group differences (all F < 0.49, i > 0.05), reinforcing their limited discriminative capacity.

The visualization details of the above results are shown in Table 2 and Figure 1. Both cohorts demonstrated AD groups showed 12-15% higher entropy values than NC/MCI (BABRI: Δentropy = 0.21; ADNI: Δentropy = 0.08), with effect sizes (η²) ranging 0.18–0.31. Despite cohort differences in baseline values (BABRI temperature: 168.02 ± 164.83; ADNI: 179.36 ± 46.35), temperature indexes consistently failed to distinguish diagnostic groups. These results highlight entropy as a potential biomarker for AD-related cortical disorganization, while temperature measures may reflect nonspecific age-related changes.

**Figure 1.**
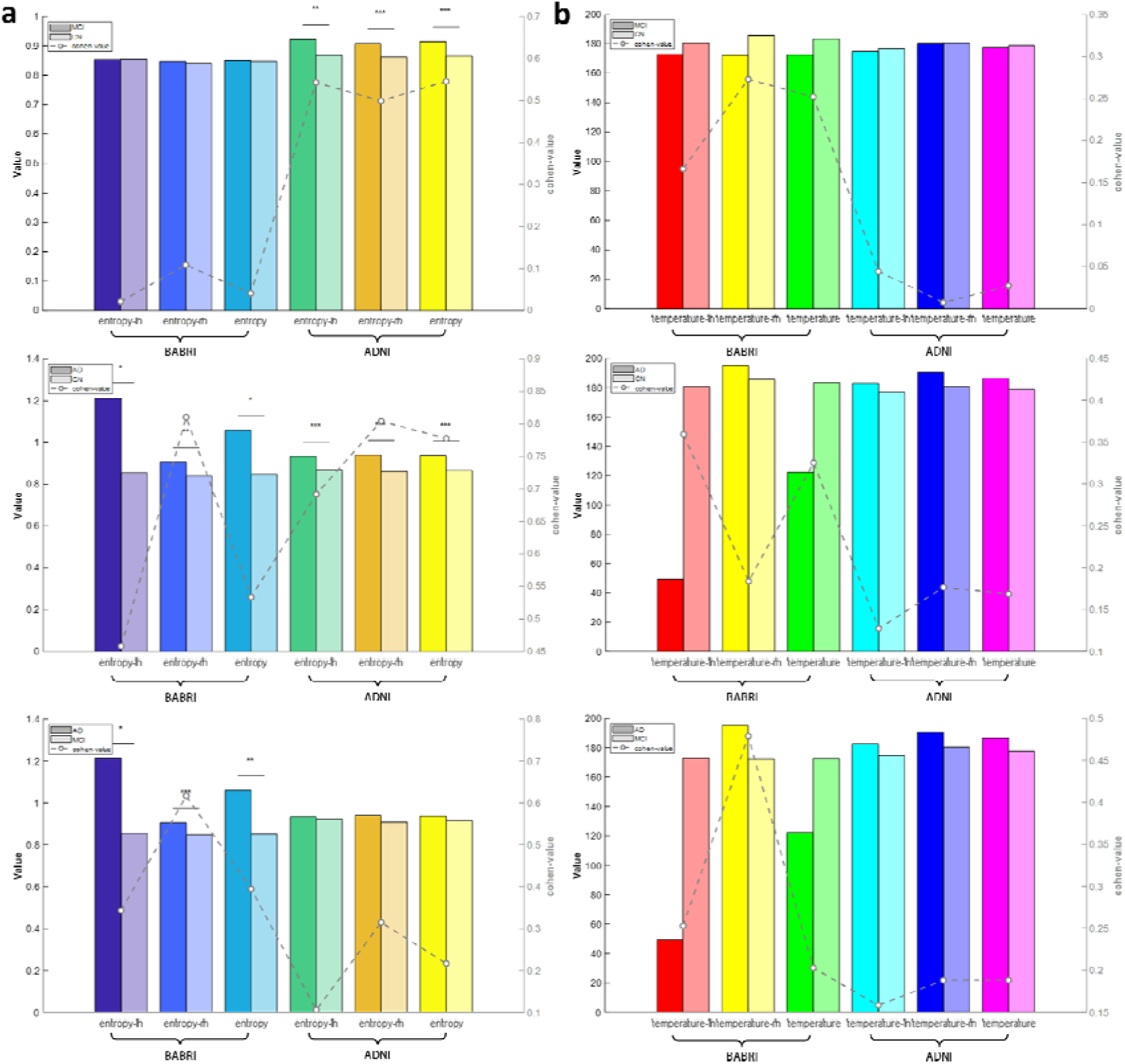
The inter-group distribution and effect intensity of entropy and temperature indicators in diagnosis. (a) Entropy values (mean ± SEM) in left hemisphere, right hemisphere, and global cortex for NC (normal control), MCI (mild cognitive impairment), and AD (Alzheimer’s disease) groups. Significant entropy elevation in AD groups versus NC/MCI is observed in both cohorts (BABRI: F = 3.07–7.64, *p*<0.05–0.001; ADNI: F = 10.77–12.21, *p*<0.001), with AD showing 12–15% higher entropy. (b) Temperature indexes show no significant intergroup differences in either cohort (all F<0.5, *p*>0.05).

**Table 2.**
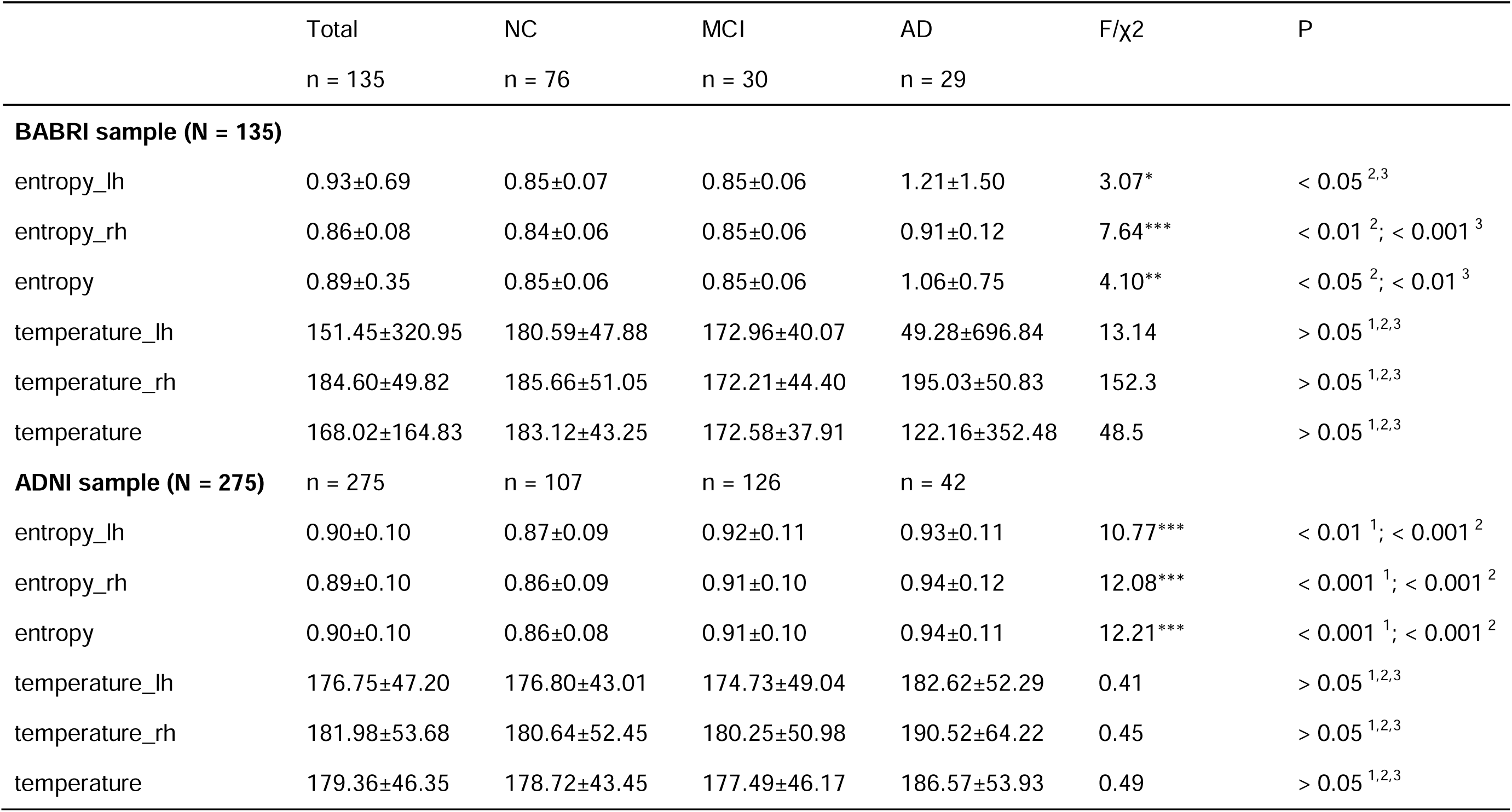
Group differences in Entropy and Temperature in BABRI and ADNI Cohorts

| | Total | NC | MCI | AD | F/$\chi^2$ | P |
| --- | --- | --- | --- | --- | --- | --- |
|  | n = 135 | n = 76 | n = 30 | n = 29 |  |  |
| **BABRI sample (N = 135)** |  |  |  |  |  |  |
| entropy_lh | 0.93±0.69 | 0.85±0.07 | 0.85±0.06 | 1.21±1.50 | 3.07* | < 0.05<sup>2,3</sup> |
| entropy_rh | 0.86±0.08 | 0.84±0.06 | 0.85±0.06 | 0.91±0.12 | 7.64*** | < 0.01<sup>2</sup>; < 0.001<sup>3</sup> |
| entropy | 0.89±0.35 | 0.85±0.06 | 0.85±0.06 | 1.06±0.75 | 4.10** | < 0.05<sup>2</sup>; < 0.01<sup>3</sup> |
| temperature_lh | 151.45±320.95 | 180.59±47.88 | 172.96±40.07 | 49.28±696.84 | 13.14 | > 0.05<sup>1,2,3</sup> |
| temperature_rh | 184.60±49.82 | 185.66±51.05 | 172.21±44.40 | 195.03±50.83 | 152.3 | > 0.05<sup>1,2,3</sup> |
| temperature | 168.02±164.83 | 183.12±43.25 | 172.58±37.91 | 122.16±352.48 | 48.5 | > 0.05<sup>1,2,3</sup> |
| **ADNI sample (N = 275)** |  |  |  |  |  |  |
|  | n = 275 | n = 107 | n = 126 | n = 42 |  |  |
| entropy_lh | 0.90±0.10 | 0.87±0.09 | 0.92±0.11 | 0.93±0.11 | 10.77*** | < 0.01<sup>1</sup>; < 0.001<sup>2</sup> |
| entropy_rh | 0.89±0.10 | 0.86±0.09 | 0.91±0.10 | 0.94±0.12 | 12.08*** | < 0.001<sup>1</sup>; < 0.001<sup>2</sup> |
| entropy | 0.90±0.10 | 0.86±0.08 | 0.91±0.10 | 0.94±0.11 | 12.21*** | < 0.001<sup>1</sup>; < 0.001<sup>2</sup> |
| temperature_lh | 176.75±47.20 | 176.80±43.01 | 174.73±49.04 | 182.62±52.29 | 0.41 | > 0.05<sup>1,2,3</sup> |
| temperature_rh | 181.98±53.68 | 180.64±52.45 | 180.25±50.98 | 190.52±64.22 | 0.45 | > 0.05<sup>1,2,3</sup> |
| temperature | 179.36±46.35 | 178.72±43.45 | 177.49±46.17 | 186.57±53.93 | 0.49 | > 0.05<sup>1,2,3</sup> |

### 3.3 Entropy Captures Heterogeneity in MCI Subtypes and Associates with Amyloid-β Accumulation

Our findings revealed that differences between CN and MCI were only observed in the ADNI dataset, while distinctions between MCI and AD emerged exclusively in the BABRI dataset. This divergence may stem from the fact that MCI populations in ADNI and BABRI represent distinct transitional phases along the CN-to-AD continuum. Specifically, MCI individuals in ADNI might be closer to the AD conversion stage, whereas those in BABRI likely remain in a more stable compensatory phase. Based on this observation, we propose a sub-hypothesis that Shannon entropy of brain activity could capture the heterogeneity within clinically defined MCI states. To test this, we stratified the MCI cohort in the ADNI dataset (N = 275) into three subgroups: reversed MCI (transitioning to CN), stable MCI (remaining MCI), and progressed MCI (transitioning to AD) (Table 3).

**Table 3.**
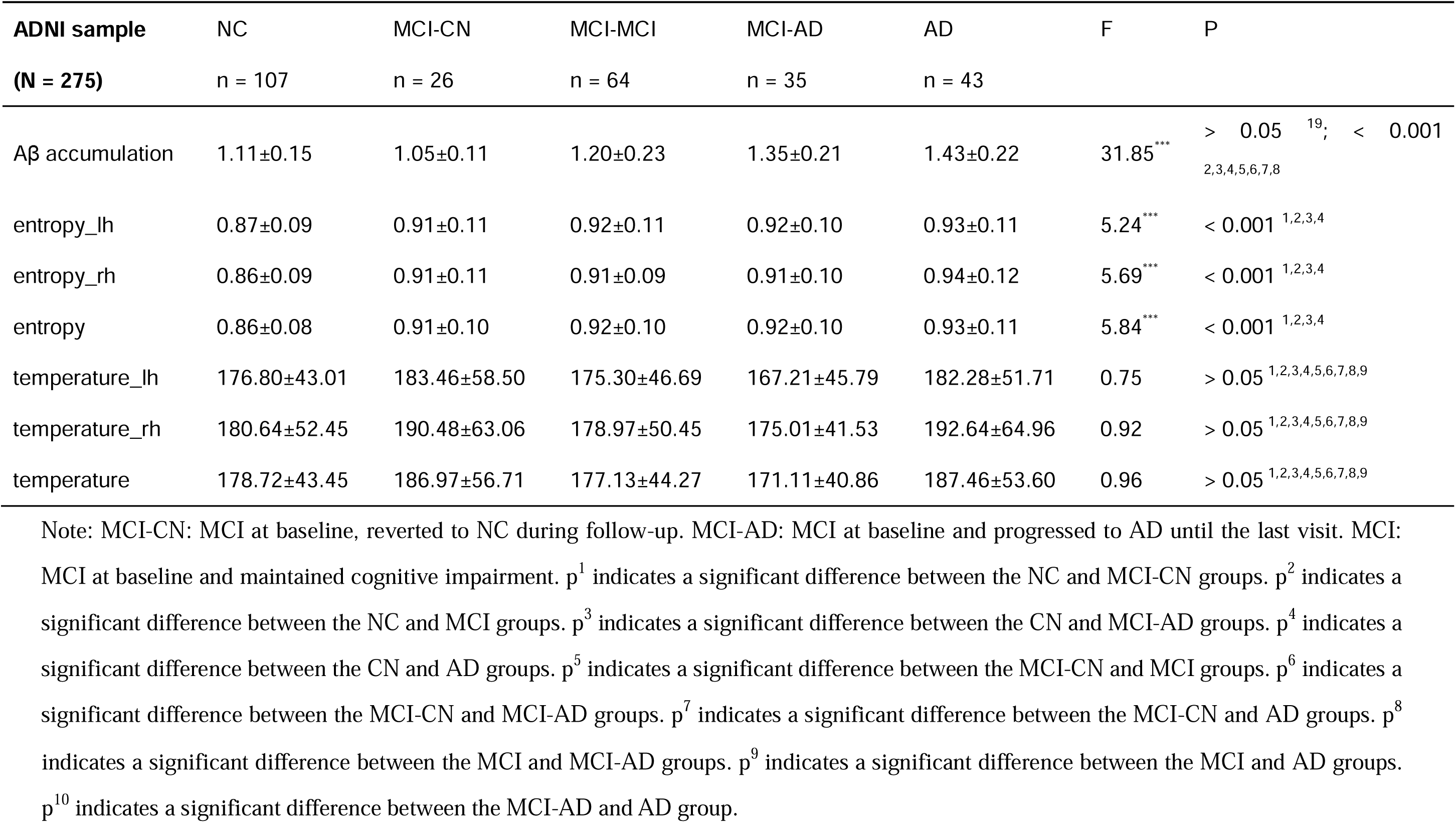
Group Differences in Entropy and Temperature Across MCI Progression Subgroups in the BABRI and ADNI Cohorts

Neurobiological characteristics exhibited significant heterogeneity across five cognitive transition groups (NC, MCI-CN, MCI-MCI, MCI-AD, AD). Amyloid-β (Aβ) accumulation demonstrated a clear pathological continuum with entropy indexes, whereas temperature parameters showed no significant group differences. Aβ levels progressively increased with disease severity (ANOVA: *F* = 31.85, *p* < 0.001), with the NC group (1.05 ± 0.11) showing significantly lower values than the MCI group (1.20 ± 0.23, *p* < 0.001), MCI-AD subgroup (1.35 ± 0.21, *p* < 0.001), and AD group (1.43 ± 0.22, *p* < 0.001). Notably, the MCI-AD subgroup exhibited higher Aβ levels compared to both MCI-CN (1.35 vs. 1.05, *p* < 0.001) and MCI-MCI subgroups (1.35 vs. 1.20, *p* < 0.001), underscoring the critical role of Aβ accumulation in AD progression.

Entropy indexes further stratified the groups (all *F* > 5.2, *p* < 0.001). Global entropy in the NC group (0.86 ± 0.08) was significantly lower than in MCI-MCI (0.92 ± 0.10, *p* < 0.001), MCI-AD (0.92 ± 0.10, *p* < 0.001), and AD groups (0.93 ± 0.11, *p* < 0.001). Right hemisphere entropy displayed a gradient increase: AD (0.94 ± 0.12) > MCI-AD (0.91 ± 0.10) > NC (0.86 ± 0.09). In contrast, brain temperature parameters showed no significant differences across groups (all *F* < 1.0, *p* > 0.05). Subsequent Spearman correlation analyses (Supplementary Figure 1) revealed significant associations between Aβ, entropy indexes, and disease progression (*p* < 0.05). Additionally, partial correlation analyses between entropy/temperature indexes and A β deposition, AVLT scores, and MMSE scores revealed that entropy indexes exhibited significant correlations with both Aβ deposition (r = 0.14, *p* < 0.05; r = 0.13, *p* < 0.05) and AVLT scores (r = −0.18, *p* < 0.001; r = −0.20, *p* < 0.001; r = −0.20, *p* < 0.001), but not with MMSE scores (r < 0.1, *p* > 0.05). In contrast, temperature indexes showed no significant associations with Aβ deposition, AVLT scores, or MMSE scores (see Supplementary Table 1 and Figure 2).

**Figure 2.**
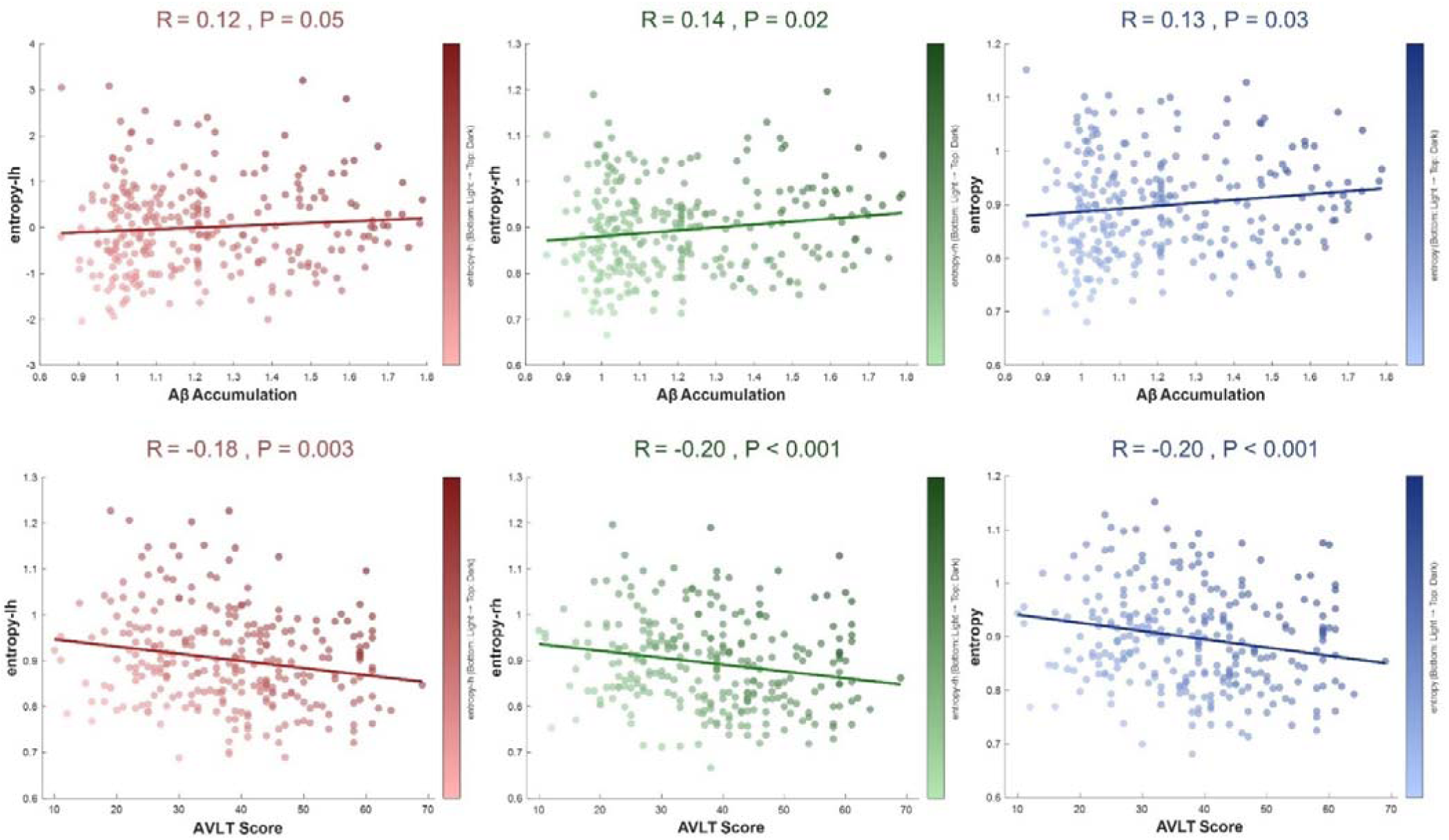
The entropy/temperature index is associated with pathology-cognition. (A) Partial correlations of entropy indexes with Aβ deposition, and AVLT (episodic memory). Entropy significantly correlates with Aβ (r = 0.13–0.14, *p* < 0.05) and AVLT (r = –0.18 to –0.20, *p* < 0.001), but not MMSE. Temperature shows no associations.

The above results indicate that entropy may serves as a robust biomarker for tracking the CN-to-AD pathological continuum.

### 3.4 Non-Linear Entropy Trajectories Link MCI to AD Conversion

We further analyzed longitudinal trends of Shannon entropy and temperature parameters across MCI subgroups during the transition process, as described in Method 2.6. Results demonstrated distinct temporal patterns: the reversed MCI exhibited a linear decline in Shannon entropy over time (Figure 3.a), while the stable MCI maintained stable entropy levels throughout the observation period (Figure 3.b). Notably, only the MCI progressor subgroup displayed a significant non-linear entropy trajectory during the transition to AD (t = 1.39, *p* < 0.01). Specifically, entropy declined from baseline to the first follow-up but plateaued in subsequent follow-ups (Figure 3.c). This biphasic trajectory may reflect the pathological progression of MCI, where initial compensatory mechanisms fail as neurodegeneration accelerates, leading to a stabilization phase marked by advanced pathological burden.

**Figure 3.**
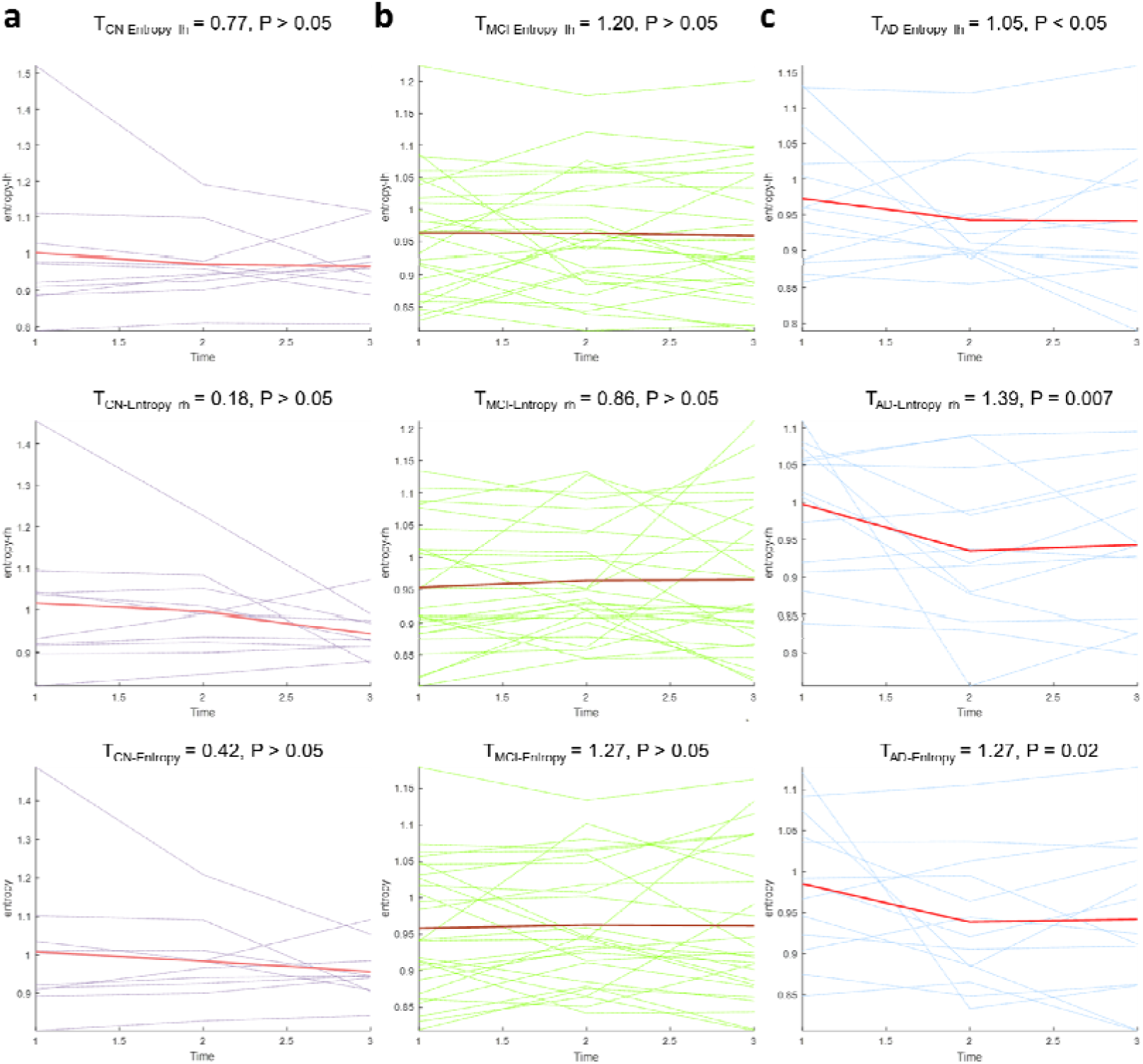
Longitudinal trajectories of entropy across MCI subtypes during cognitive transition: (A) Reversed MCI (transitioning to CN) shows linear entropy decline over time, indicating functional normalization; (B) Stable MCI maintains consistent entropy levels, reflecting compensatory equilibrium; (C) MCI progressors (transitioning to AD) exhibit a biphasic nonlinear trajectory—initial entropy decline (baseline → first follow-up) followed by plateauing (subsequent follow-ups) (t = 1.39, *p* < 0.01), marking the transition from compensatory effort to pathological stabilization. Shaded areas denote 95% confidence intervals. Temperature trajectories (not shown) displayed no significant trends across subgroups.

### 3.5 Entropy Predicts Aβ Deposition and Distinguishes Neurodegenerative Stages

The results demonstrated that only Shannon entropy exhibited significant differences across CN, MCI, and AD groups. We therefore constructed linear regression and support vector machine (SVM) models under leave-one-out cross-validation (see Methods). The linear regression model aimed to predict Aβ deposition using entropy indexes, while the SVM model aimed to distinguish normal aging from pathological decline.

ROC curve analysis of the SVM classifier revealed the discriminative power of entropy indexes for cognitive impairment subgroups (Figure 4a-c). Left-hemisphere entropy (entropy-lh) consistently outperformed right-hemisphere and global entropy across all classification tasks. Specifically, in CN vs. MCI classification, entropy-lh achieved the highest AUC (0.901), significantly surpassing global entropy (AUC = 0.498) and Aβ (AUC = 0.539). Similarly, in CN vs. AD discrimination, entropy-lh (AUC = 0.873) exceeded right-hemisphere entropy (AUC = 0.519) and other biomarkers. For MCI vs. AD classification, entropy-lh (AUC = 0.724) also outperformed other biomarkers (all AUCs ≤ 0.501). Notably, Aβ showed limited discriminative capacity across all tasks (AUC = 0.485–0.539), likely due to sample heterogeneity, whereas entropy-lh maintained robust performance in CN-MCI (AUC = 0.901) and CN-AD (AUC = 0.873) classifications, highlighting its potential as a neurodegenerative biomarker.

**Figure 4.**
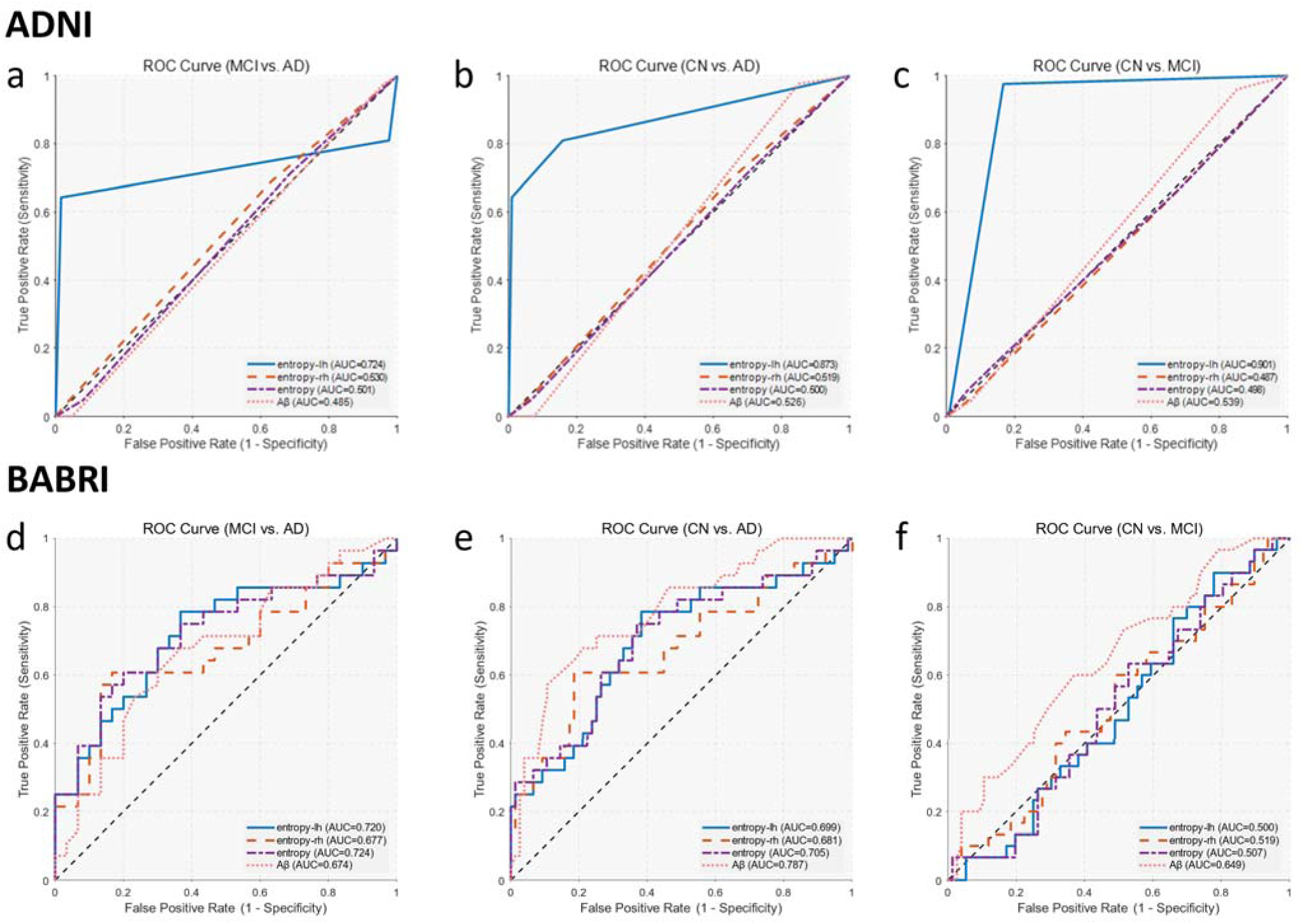
Diagnostic classification performance of entropy indexes in both cohorts across different progression stages. ADNI: (a) For MCI vs. AD classification, left-hemisphere entropy (entropy-lh) achieved the highest performance (AUC = 0.724), outperforming right-hemisphere entropy (AUC = 0.530), global entropy (AUC = 0.501), and Aβ (AUC = 0.485); (b) In CN vs. AD classification, entropy-lh again showed superior discriminative power (AUC = 0.873), while other indexes approached chance level (entropy-rh = 0.519, global entropy = 0.500, Aβ = 0.526). (c) For CN vs. MCI classification, entropy-lh yielded excellent performance (AUC = 0.901), substantially surpassing right-hemisphere entropy (AUC = 0.487), global entropy (AUC = 0.498), and Aβ (AUC = 0.539). **BABRI:** (d) In MCI vs. AD classification, all entropy indexes—left-hemisphere (AUC = 0.720), right-hemisphere (AUC = 0.677), and global entropy (AUC = 0.724)—showed comparable performance to Aβ (AUC = 0.674); (e) In CN vs. AD discrimination, global entropy achieved the best entropy-based performance (AUC = 0.705), while Aβ outperformed all entropy indexes (AUC = 0.787); (f) In CN vs. MCI classification, entropy indexes were near chance level (AUCs = 0.500–0.519), whereas Aβ showed modest discrimination (AUC = 0.649).

We also evaluated their diagnostic utility in the BABRI cohort (Figure 4d–f). In distinguishing MCI from AD (Figure 4d), all three entropy indexes—left-hemisphere (AUC = 0.720), right-hemisphere (AUC = 0.677), and global entropy (AUC = 0.724) —demonstrated moderate classification performance, on par with Aβ (AUC = 0.674).

This suggests that entropy measures are comparably sensitive to Aβ in capturing disease progression from MCI to AD. In CN vs. AD classification (Figure 4e), global entropy achieved the highest entropy-based AUC (0.705), slightly outperforming left-hemisphere (AUC = 0.699) and right-hemisphere (AUC = 0.681). However, Aβ (AUC = 0.787) showed stronger performance in this task, indicating greater separability at the extreme ends of the disease spectrum. In contrast, entropy indexes performed poorly in CN vs. MCI discrimination (Figure 4f), with AUCs near chance level (entropy-lh = 0.500, entropy-rh = 0.519, global entropy = 0.507), while Aβ showed modest discriminability (AUC = 0.649). These findings suggest that entropy is less sensitive to early-stage, preclinical changes in this cohort, potentially due to limited signal in structural-functional disorganization at this early stage.

In generalized linear models predicting Aβ deposition, only right-hemisphere entropy (RMSE = 0.22, 95% CI [0.21, 0.24]) and global entropy (RMSE = 0.22, 95% CI [0.21, 0.24]) exhibited lower prediction errors compared to the baseline model (RMSE = 0.38, 95% CI [0.35, 0.41]), corresponding to a 42.1% reduction in error (ΔRMSE = −0.16, P_permutation_ < 0.001). Similar trends were observed in the BABRI cohort, where right-hemisphere entropy (RMSE = 0.13, 95% CI [0.11, 0.14]) and global entropy (RMSE = 0.12, 95% CI [0.11, 0.14]) reduced errors by 38.7% (ΔRMSE = −0.12, P_permutation_ = 0.002) relative to the baseline (RMSE = 0.31, 95% CI [0.27, 0.35]). These findings underscore the dual utility of entropy indexes: left-hemisphere entropy excels in diagnostic classification, while right-hemisphere and global entropy robustly predict Aβ burden, collectively positioning entropy as a versatile biomarker in neurodegenerative research.

## 4 Discussion

This study, through validation in two independent cohorts, is the first to systematically demonstrate the diagnostic value and pathological relevance of brain activity entropy—quantified by Shannon entropy based on gray matter intrinsic modes—as a novel biomarker for Alzheimer’s disease (AD).

Our findings reveal that both whole-brain and hemispheric entropy values are significantly elevated in AD patients compared to cognitively normal (CN) individuals and those with mild cognitive impairment (MCI), with the left hemisphere showing the highest diagnostic accuracy. This result supports the sensitivity of entropy in quantifying disruptions in cortical information integration. Drachman’s (2006) core proposition—that AD represents an accelerated stage of entropy increase associated with brain aging—provides the theoretical foundation for our analysis[51]. Our entropy index appears to quantify the degree of disorder in neural information integration. In normal aging, entropy increases gradually, reflecting reduced neuronal redundancy and the depletion of cognitive reserve[52, 53]. However, in the pathological aging associated with AD, entropy increases disproportionately, potentially due to Aβ-induced ion channel dysfunction that renders the gray matter microenvironment chaotic[54], ultimately disrupting the functional mapping of intrinsic modes. Moreover, Drachman emphasized the nonlinear relationship between AD neuropathology and cognitive impairment. Our findings offer a novel interpretation: entropy may reflect the buffering role of cognitive reserve. Under equivalent Aβ burden, individuals with lower entropy may delay the onset of dementia through functional reorganization (e.g., compensatory cortical recruitment) [55]. Conversely, when Aβ exceeds a critical threshold, a transition to pathological aging may occur, characterized by the collapse of geometric constraints in gray matter.

This breakdown leads to a significant increase in systemic disorder and eventual functional disintegration of the brain[56, 57].

In contrast, functional reconfiguration temperature did not show significant group differences in either cohort, suggesting that it may primarily reflect non-specific aging processes (e.g., global declines in brain network efficiency) rather than AD-specific pathology[58, 59]. This discrepancy may be attributed to fundamental differences in the computational principles underlying the two indexes. Entropy is derived by computing the inner product between each vertex’s functional neural time series (fMRI BOLD signals) and structural intrinsic modes—represented by Laplacian eigenvectors constrained by gray matter geometry. This operation yields vertex-specific β weights, from which the entropy of their distribution is calculated. The inner product preserves the physical constraints imposed by gray matter geometry on local neural dynamics[60], allowing entropy to directly reflect the degree of chaos in microscale circuit-level information integration. In contrast, the temperature index aggregates vertex data into macroscale brain regions (e.g., 400 parcels based on the Schaefer 2018 atlas), which sacrifices vertex-level variability. Moreover, it uses eigenmode-based modeling to approximate the macroscale functional connectivity (FC) matrix, capturing only linear regional correlations while ignoring nonlinear dynamics[61]. Therefore, entropy is more sensitive to AD-specific spatiotemporal dynamics at the microscale level[62], whereas temperature, being dependent on FC-based pathways, tends to lose disease specificity at the macroscale. Consequently, entropy emerges as a superior biomarker for the ultra-early detection of AD, while temperature may be more appropriate for assessing normative aging processes.

Entropy reveals the heterogeneity of MCI and predicts Aβ deposition. Specifically, entropy effectively stratifies MCI subtypes in the ADNI cohort by distinguishing their conversion trajectories. Progressive MCI (MCI-AD) exhibited significantly higher entropy values and Aβ burden (1.35C±C0.21) compared to stable (MCI-MCI: 1.20C±C0.23) and reverting (MCI-CN: 1.05C±C0.11) subtypes, supporting the utility of entropy in capturing the continuous evolution of preclinical pathology.

Longitudinal analysis further revealed a nonlinear trajectory of entropy during MCI-to-AD conversion, characterized by a biphasic pattern: an initial compensatory phase with a transient decline in entropy, followed by a decompensatory phase where entropy plateaus. This dynamic profile offers a quantifiable window to predict the critical point of conversion. In the early phase, the disruption caused by Aβ plaques impairs neuronal plasticity and reduces the efficiency of information transfer between neurons[63, 64]. To maintain overall brain function, the network responds with compensatory redundant connectivity, which temporarily reduces entropy by simplifying network complexity and offsetting localized damage[65]. As the disease progresses, the plateau phase in entropy is likely driven by widespread neurodegeneration. At this stage, synaptic loss exceeds 30%, and gray matter geometry is substantially disrupted, marking the collapse of compensatory mechanisms and the onset of functional failure[66].

Left hemispheric entropy plays a dominant role in diagnostic classification. It achieved the best performance in distinguishing cognitively normal (CN) individuals from Alzheimer’s disease (AD) patients, with an AUC of 0.901. This may reflect the vulnerability of key regions in the left hemisphere, which are responsible for language and logical integration. Regions such as hubs of the default mode network[67] are particularly susceptible to Aβ deposition and tau tangles in AD. The elevated entropy in these areas suggests that left hemisphere entropy is sensitive to structural damage and functional decline. It may help identify meaningful prodromal symptoms of AD earlier than traditional markers. Right hemispheric entropy, on the other hand, is more closely associated with Aβ pathology prediction. The right hemisphere supports spatial navigation and episodic memory, functions that are often affected in the early stages of AD[52]. Abnormal entropy in this hemisphere may reflect early molecular changes. Notably, the right posterior cingulate cortex is one of the initial sites for Aβ accumulation[68]. Entropy increases in this area may indicate impaired clearance mechanisms, possibly due to dysfunction in internal systems like the glymphatic pathway[69]. Thus, right hemisphere entropy could serve as a sensitive marker of early molecular pathology, even before cognitive symptoms become apparent.

## 5 Limitation

Despite the strong discriminative power of entropy indexes demonstrated across cohorts, several limitations should be acknowledged.

First, pathological validation remains incomplete. Although entropy showed a modest correlation with Aβ-PET (r = 0.14), tau-PET and cerebrospinal fluid (CSF) biomarkers were not included in the analysis. Since tau pathology is more closely linked to cognitive decline than Aβ, future studies should assess whether entropy can also capture tau-related network disintegration.

Second, MCI subtyping was based solely on clinical progression categories (MCI-CN, MCI-MCI, MCI-AD), without incorporating biomarker-defined classifications such as A+T+ or A−T−. This limitation may obscure distinct entropy trajectories in biologically diverse MCI subtypes, such as limbic-predominant versus hippocampal-sparing AD.

Third, the lack of significant group differences in the temperature index (all p > 0.05) may reflect methodological constraints. Parcel-based smoothing of functional connectivity likely diluted vertex-level dynamics that are critical for detecting AD-related changes. Alternative formulations of temperature, such as vertex-wise or spectral methods, have not yet been explored and may offer improved sensitivity.

Fourth, the biphasic entropy trajectory observed in MCI converters was based on a limited number of follow-up time points, typically only two after baseline. Higher-frequency longitudinal sampling is necessary to accurately identify the inflection point between early compensatory decline and the later pathological plateau. This would help refine the timing of potential interventions.

Fifth, the relationship between entropy and cognition appears to be domain-specific. Entropy correlated with episodic memory performance, as measured by the AVLT, but not with global cognition assessed by the MMSE. Including more detailed cognitive assessments, such as tests of executive function or visuospatial abilities, may clarify how entropy changes relate to specific cognitive domains across brain hemispheres.

## 6 Conclusion

This multi-cohort study establishes Shannon entropy as a robust, dual-purpose biomarker for Alzheimer’s disease (AD) pathology, demonstrating significant efficacy in both diagnostic classification and pathological prediction. Entropy index consistently differentiated AD from NC and MCI across BABRI and ADNI cohorts, with left-hemisphere entropy being the optimal classifier for neurodegenerative staging and right-hemisphere entropy dominating Aβ burden prediction. Moreover, entropy was able to stratify clinically defined MCI into biologically distinct subtypes and revealed a biphasic entropy shift in MCI→AD converters. The hemispheric asymmetry in entropy utility offers a unified framework for "structure-function decoupling" in association cortices. Entropy’s superiority over Aβ-PET and resistance to demographic confounds make it a cost-effective, non-invasive tool for early risk stratification and identifying the "inflection point" in MCI biphasic trajectories for precision neuromodulation.

## Data Availability

All data produced in the present study are available upon reasonable request to the authors.

## Acknowledgments and Funding Disclosure

This work was supported by STI2030-Major Projects (2022ZD0211600), the Natural Science Foundation of China (Grants No. 32171085), the National Natural Science Foundation of China (Grants No. 12405062), State Key Program of National Natural Science of China (Grants No. 82130118), the Fundamental Research Funds for the Central Public welfare research institution (ZZ13-YQ-073, Z0601), Open Research Fund of the State Key Laboratory of Cognitive Neuroscience and Learning and Tang Scholar.

## Data Availability

The data sets analyzed during the study are available from the corresponding author upon reasonable request.

