## Supplemental tables for "Shannon Entropy of Gray Matter Eigenmodes: A Novel Biomarker for Alzheimer’s Disease and Heterogeneous MCI Trajectories"

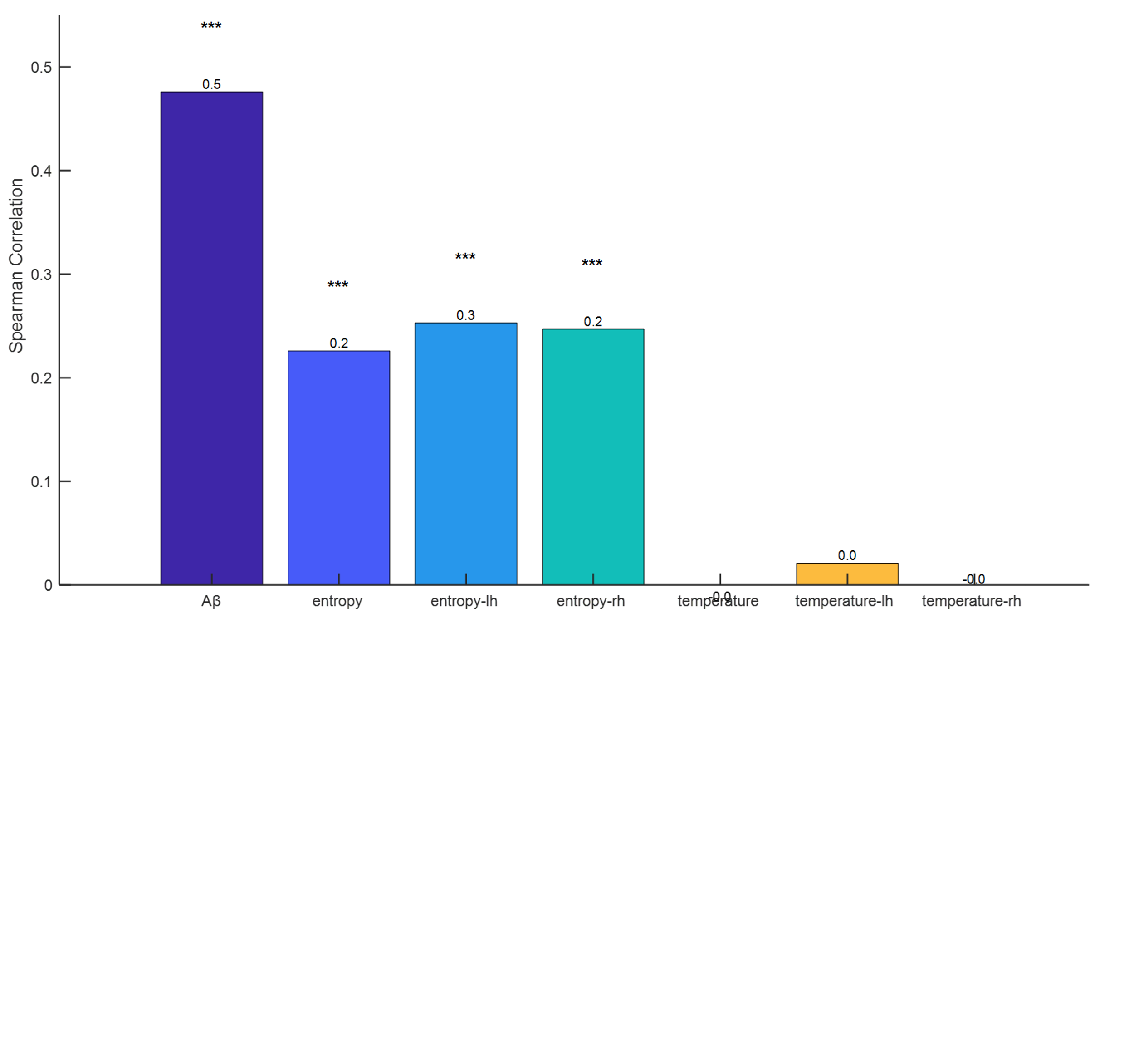

**Supplementary Figure 1. The cross-group correlation between entropy and Aβ deposition.** Spearman correlations between Aβ deposition and entropy metrics across cognitive transition groups (NC, MCI-CN, MCI-MCI, MCI-AD, AD). Temperature metrics display no significant trends.

**Supplementary Table 1** Correlation of entropy and temperature with Aβ accumulation and cognitive performance

|  | e_lh | e_rh | entropy | t_lh | t_rh | temper | Total | MMSE | AVLT |
| --- | --- | --- | --- | --- | --- | --- | --- | --- | --- |
| e_lh | -- | 0.85^***^ | 0.96^***^ | 0.12 | 0.08 | 0.10 | 0.08 | -0.04 | -0.18^***^ |
| e_rh | 0.85^***^ | -- | 0.96^***^ | 0.03 | 0.00 | 0.02 | 0.14^*^ | -0.08 | -0.20^***^ |
| entropy | 0.96^***^ | 0.96^***^ | -- | 0.08 | 0.04 | 0.06 | 0.13^*^ | -0.06 | -0.20^***^ |
| t_lh | 0.12 | 0.03 | 0.08 | -- | 0.68^***^ | 0.91^***^ | 0.05 | -0.05 | -0.06 |
| t_rh | 0.08 | 0.00 | 0.04 | 0.68^***^ | -- | 0.93^***^ | 0.05 | -0.07 | -0.09 |
| temper | 0.10 | 0.02 | 0.06 | 0.91^***^ | 0.93^***^ | -- | 0.05 | -0.07 | -0.08 |
| Aβ | 0.12 | 0.14^*^ | 0.13^*^ | 0.05 | 0.05 | 0.05 | -- | -0.39^***^ | -0.41^***^ |
| MMSE | -0.04 | -0.08 | -0.06 | -0.05 | -0.07 | -0.07 | -0.39^***^ | -- | 0.59^***^ |
| AVLT | -0.18^***^ | -0.20^***^ | -0.20^***^ | -0.06 | -0.09 | -0.08 | -0.41^***^ | 0.59^***^ | -- |

Note: Pearson correlation coefficients (*r*) are reported; *p*-values in parentheses: ** < .05,* ******** *< .01, ***** < .001*; Diagonal entries = 1.00 (variable self-correlation); Bonferroni correction applied for multiple comparisons
